# Completeness and Quality of Neurology Referral Letters Generated by a Large Language Model for Standardized Scenarios

**DOI:** 10.1101/2025.06.12.25329503

**Authors:** Watcharasarn Rattananan

**Author notes:** Corresponding author: Watcharasarn Rattananan, M.D., M.B.A. Correspondence, Address: 492/1 Rama I Rd, Pathum Wan, Bangkok 10330, Thailand. Clinical trial number: not applicable.

## Abstract

**Background:** Large Language Models (LLMs) offer promising applications in healthcare, including drafting referral letters. However, access to LLMs specifically designed for medical practice remains limited. While ChatGPT is widely available, its ability to generate comprehensive and clinically appropriate neurology referral letters remains uncertain. This study aimed to systematically evaluate the completeness and quality of neurology referral letters generated by ChatGPT for standardized clinical scenarios.

**Methods:** Five standardized clinical scenarios representing common neurological complaints (e.g., headache, memory problems, stroke/TIA, tremor, radiculopathy) were used. ChatGPT generated 10 referral letters per scenario using the same prompt. Each letter was evaluated using a predefined scoring rubric assessing for completeness (demographics, chief complaint, history of present illness, physical exam findings, management, and consultation questions), and quality (language level, structure, and letter length), reviewed by a physician dual-board-certified in neurology and family medicine.

**Results:** ChatGPT reliably included key elements, achieving an average completeness score of 87%. The output demonstrated high language appropriateness (average scores of 92% for language level and 90% for structure). Variability was noted in case management (mean score of 2.18, standard deviation of 0.85). Noticeable gaps in content, particularly in history of present illness and physical findings, were identified in 72% (36/50) of the letters. Document length (250–450 words) was acceptable, though word count often exceeded expectations.

**Conclusions:** ChatGPT demonstrated high efficiency and utility in generating neurology referral letters but required physician oversight to address variability in case management and minor content gaps. Access to tailored LLMs trained for medical documentation could improve outcomes while safeguarding patient privacy.

**Trial registration:** Not required (no intervention on human participants)

## Background

Advances in artificial intelligence (AI) have introduced transformative tools in healthcare, with earlier applications focusing on diagnosis, treatment and prognosis (Ali, 2020; Jiang, 2017). Recently, Large Language Models (LLMs) emerged as potential aids in clinical documentation (Liu et al., 2024) or scribing (Avant, 2024; Shah, 2025). Referral letters play a pivotal role in patient care coordination, bridging primary and specialty care. Accurate and complete referral letters are critical for effective communication between healthcare providers, particularly in neurology. For neurological patients, where complex patient presentations often necessitate detailed documentation, a well-crafted referral letter could significantly impact patient outcomes. This group of patients seem to need more specialized medical LLM tailored for neurological scenarios (Hillis, 2022; Marvix.ai, 2025; Voigtlaender, 2024). Unfortunately, access to medically specialized LLMs, especially for neurology, is not globally available.

ChatGPT, a prominent LLM, has been widely adopted across industries for text generation. Its potential for medical applications, including generating medical notes (Kernberg, 2024), discharge summary (Schwieger, 2024), letter (Tung, 2024), responses to patients’ questions (Ayers, 2023). Utilizing other LLMs in drafting physicians’ letters (Harris, 2024; MEDICA Trade Fair, 2024) has been sparked considerable interest as well. However, unlike tasks such as summarizing patient records or generating discharge instructions, referral letters demand different structure, contextual understanding, and adherence to professional standards. Although LLMs have demonstrated proficiency in generating coherent text, their ability to address the needs of medical communication remains understudied. Most existing studies focus on broader applications, such as summarizing medical notes (Heilmeyer, 2024) or extracting information from electronic health records. This study narrows its scope to assess the specific task of generating referral letters, a critical component of interdisciplinary medical communication between primary physicians and neurologists. By examining the performance of ChatGPT in standardized neurological scenarios, we aim to quantify the completeness and quality of AI-generated referral letters, identify common content gaps and areas for improvement, as well as discuss implications for clinical practice.

## Methods

### Study Design

Five standardized scenarios representing common neurological complaints were developed to mimic real-world neurological consultations. These included:

**Case 1**: A 32-year-old female with chronic daily headaches, likely migraines, complicated by medication overuse.

**Case 2**: A 68-year-old female presenting with memory problems, concerning for early dementia.

**Case 3**: A 68-year-old male with transient ischemic attack (TIA) symptoms.

**Case 4**: A 68-year-old female with tremors suggestive of Parkinson’s disease.

**Case 5:** A 58-year-old male with right-sided radiculopathy consistent with L5-S1 involvement.

Each scenario provided essential clinical details, including history, physical findings, and initial assessments. Emphasis on different angles of case information was applied depending on case types. For instance, a case with memory problem provided relatively more detail in history of present illness part, while a case with tremor or radiculopathy showed more physical exam findings. Combining the five cases, all parts of analyzed medical information should be well balanced. ChatGPT (GPT-4o) was prompted to generate 10 referral letters per scenario, one-by-one, using a consistent instruction to ensure consistency in inputs. The generated letters were reviewed for completeness and quality, ensuring relevance to each scenario.

### Scoring and Analysis

A predefined rubric (Table 1), designed by expert input, comprised of the following items:

- *Completeness* (20 points): Inclusion of demographics, chief complaint (CC), history of present illness (HPI), physical exam findings (PE), management (Mx), and consultation questions (Q). These categories reflect the essential components of a neurology referral letter.
- *Quality* (10 points): Language level, structure, and adherence to length guidelines. Criteria were adapted to align with clinical standards, emphasizing clarity, professionalism, and conciseness.

**Table 1:**
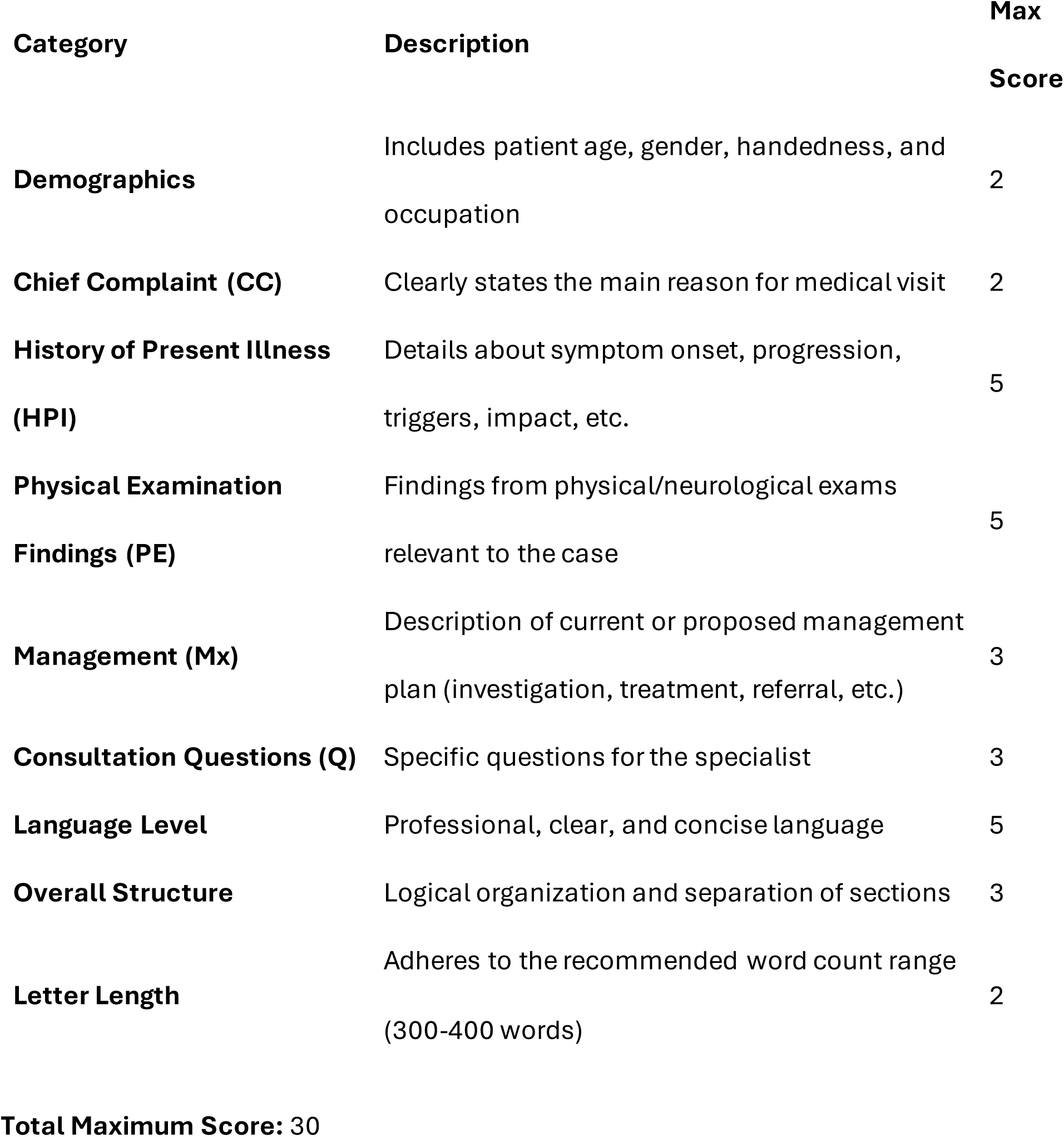
Rubric for Evaluation of Neurology Referral Letters.

| Category | Description | Max Score |
| --- | --- | --- |
| Demographics | Includes patient age, gender, handedness, and occupation | 2 |
| Chief Complaint (CC) | Clearly states the main reason for medical visit | 2 |
| History of Present Illness (HPI) | Details about symptom onset, progression, triggers, impact, etc. | 5 |
| Physical Examination Findings (PE) | Findings from physical/neurological exams relevant to the case | 5 |
| Management (Mx) | Description of current or proposed management plan (investigation, treatment, referral, etc.) | 3 |
| Consultation Questions (Q) | Specific questions for the specialist | 3 |
| Language Level | Professional, clear, and concise language | 5 |
| Overall Structure | Logical organization and separation of sections | 3 |
| Letter Length | Adheres to the recommended word count range (300-400 words) | 2 |

Each letter was scored by a physician dual-board-certified in neurology and family medicine. Descriptive statistics, including mean, standard deviation (SD), and percentage completion, were calculated for each rubric category. Qualitative analysis identified recurring omissions and inconsistencies. *Language appropriateness* was determined using sum scores from ‘Language level’ and ‘structure’, while *gaps in content* included scores from demographics, chief complaint, HPI and PE parts.

### Ethical Considerations

All scenarios were AI-generated to avoid patient confidentiality concerns. This ensured ethical compliance and eliminated real-world risks associated with data misuse.

## Results

### Overall Performance

The mean total score across 50 letters was 25.76/30 (85.87%), with high scores in demographic inclusion (96%), chief complaint clarity (95%), and language level (91.6%). However, content gaps were prevalent (36/50; 76%), particularly in HPI (88.4%) and PE (84.8%). The overall scores ranged from 10 – 30 indicating that the LLM-generated letters can be vastly different (Appendix A); however, given low SD (3.43), most letters are considered acceptable.

### Letter Completeness

Referral letters produced by ChatGPT demonstrated consistent inclusion of fundamental elements such as patient demographics and chief complaints (Table 2). However, significant variations in completeness were observed across different sections. The most frequently omitted details were found in the history of present illness (HPI) and physical examination (PE) components. While most letters provided a reasonable summary of the clinical scenario, they frequently omitted important negative findings. For instance, in the memory loss scenario, letters often omitted normal orientation function, while in other neurological cases, descriptions about negative family history of neurological disorders were inconsistently included.

**Table 2:**
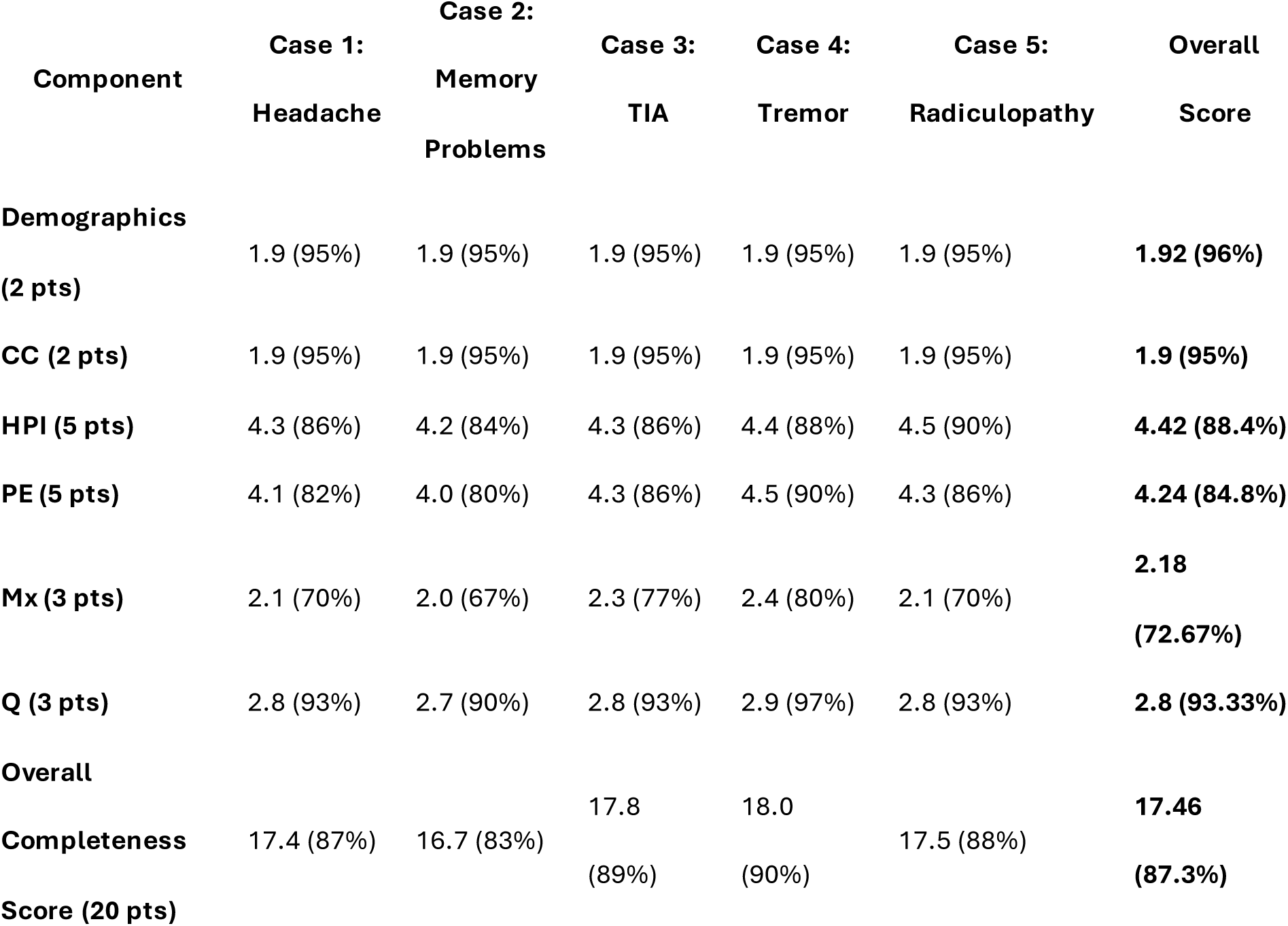
Summary of Completeness Scores for Each Case.

The management section, mostly derivative of LLM summarization, exhibited the lowest performance (scored 72.67%), with some letters failing to provide detailed differential diagnoses or clear referral justifications. Case management discussions varied in depth, with some letters offering comprehensive assessments while others provided only minimal information. Some letters described a comprehensive assessment of alternative diagnoses (e.g., differentiating between TIA and migraine aura), while others failed to mention essential considerations.

Additionally, a notable inconsistency was observed in the *consultation questions*, which are critical for guiding specialist evaluations. While some letters provided direct and well-formulated referral questions (e.g., “Would DaTscan be beneficial in ruling out Parkinson’s disease?”), others lacked specificity.

Despite these limitations, most letters met a minimum threshold of completeness sufficient for specialist review.

### Letter Quality

The generated referral letters exhibited high linguistic fluency and structural coherence, scoring an average of 91.6% for language appropriateness and 90% for structural organization (Table 3). The language used was consistently professional, concise, and grammatically accurate, making the letters readable and appropriate for medical communication. However, letter length was inconsistent, with 49% of letters either exceeding or falling short of the optimal word range. While structural organization remained strong, some letters lacked logical sequencing of information, particularly when presenting neurological examination findings or treatment plans. Furthermore, redundancies and extraneous wordings were observed in a subset of letters. Some letters repeated basic information without summarize assessment of the cases, reducing their clinical usefulness.

**Table 3:** Quality Metrics (Language, Structure, Letter Length)

| Quality Metric | Case 1:<br>Headache | Case 2:<br>Memory<br>Problems | Case 3:<br>TIA | Case 4:<br>Tremor | Case 5:<br>Radiculopathy | Overall Score |
| --- | --- | --- | --- | --- | --- | --- |
| Language<br>Level (5 pts) | 4.6 (92%) | 4.5 (90%) | 4.7 (94%) | 4.6 (92%) | 4.5 (90%) | 4.58 (91.6%) |
| Structure &<br>Organization<br>(3 pts) | 2.7 (90%) | 2.6 (87%) | 2.8 (93%) | 2.9 (97%) | 2.7 (90%) | 2.7 (90%) |
| Letter Length<br>Compliance<br>(2 pts) | 1.0 (50%) | 1.1 (55%) | 1.2 (60%) | 1.0 (50%) | 1.0 (50%) | 1.02 (51%) |
| Total Quality<br>Score (10 pts) | 8.3 (83%) | 8.2 (82%) | 8.7 (87%) | 8.5 (85%) | 8.2 (82%) | 8.38 (83.8%) |

Overall, while AI-generated letters were grammatically sound and relatively well-structured, they required manual adjustments to improve clarity, organization, and case-specific relevance before clinical use.

## Discussion

ChatGPT’s ability to produce referral letters with high completeness and quality demonstrates its potential as a time-efficient tool for clinicians. By automating routine documentation tasks, LLMs could reduce administrative burdens and enhance clinician focus on patient care. However, variability and content gaps highlight the need for physician oversight. Particularly in neurology, where diagnostic accuracy hinges on detailed documentation, these gaps could hinder care coordination.

In developing and underdeveloped countries, the shortage of medical professionals and resources poses challenges to healthcare delivery. Inaccessibility to LLM applications specifically designed for medical practice is not uncommon. In addition, language barrier can be an important obstacle in implementing LLMs in medical service infrastructure of some countries. This is due to localized training data and tailored algorithms are essential to ensure cultural and linguistic relevance. LLMs like ChatGPT and other closed- or open-source AIs could alleviate documentation burdens and enhance communication in these settings.

One of the study’s primary strengths is the use of standardized clinical scenarios, which allows for controlled assessment and direct comparability across multiple AI-generated outputs. The evaluation framework, grounded in structured rubrics assessing completeness and quality, ensures that key aspects of referral communication are systematically analyzed. Additionally, the inclusion of multiple cases covering common neurological presentations enhances the study’s generalizability to various subspecialty referrals.

Despite of these, certain limitations warrant consideration. While standardized scenarios facilitate objective assessment, they may not fully capture the complexity and variability of real-world clinical encounters, where patient narratives are often different and require adaptive reasoning. Additionally, the study focuses exclusively on English-language outputs, limiting its applicability to multilingual healthcare settings where LLMs may exhibit different performance based on linguistic and cultural environment. Therefore, although the AI-generated letters demonstrated strong structural consistency, gaps in clinical reasoning and differential diagnosis formulation suggest the need for continued physician oversight when integrating AI tools into referral workflows.

Expanding the study to include multilingual scenarios, such as Thai, Arabic, Japanese, etc., would provide valuable insights into LLM performance across languages. Incorporating real-world clinical data and feedback from end-users (clinicians and specialists) would also promote understanding and development in the AI-assisted workflow. Since LLMs’ performance is dynamic and may change overtime (Chen, 2024), it is important to collaborate with AI developers to create affordable domain-specific models which could optimize outcomes while safeguard patient privacy. Appropriate integration of such AI applications into electronic health records (EHRs) is likely to reduce physicians’ workload and burnout (Baughman, 2024; Garcia, 2024) as well as promote attitudes towards AI utilization (Allen, 2024).

### Conclusions

This study demonstrates that ChatGPT can generate neurologically focused referral letters with high completeness and quality. However, physician oversight remains crucial to address content gaps and variability. Tailored LLMs designed for medical applications could further enhance clinical communication, particularly in resource-constrained settings. Future efforts should prioritize multilingual adaptations, real-world validation, seamless integration into clinical workflows, as well as accessibility to such AI applications in underdeveloped countries.

## Data Availability

All data produced in the present study are available upon reasonable request to the authors

## List of abbreviations

AI: Artificial intelligence
CC: Chief complaint
DaT: Dopamine Transporter
EHR: Electronic health record
HPI: History of present illness
LLM: Large Language Model
Mx: Management
PE: Physical exam findings
Q: Consultation questions
SD: Standard deviation
TIA: Transient ischemic attack

## Declarations

### Ethics approval and consent to participate

Not applicable

### Consent for publication

Not applicable

### Availability of data and materials

The datasets used and/or analyzed during the current study are available from the corresponding author on reasonable request.

### Authors’ contributions

WR is the sole investigator and author of this research, responsible for study conception, data collection, analysis, manuscript drafting, and final approval of the submitted version.

## Acknowledgements

Not applicable

## Authors’ information (optional)

Not applicable

## Clinical trial number

not applicable.

## Appendix A: Sample AI-Generated Neurology Referral Letters

### A.1: High-Quality Referral Letter (TIA Case – Letter 3.10, Score: 30/30)

**[Referring Physician’s Name]**

Family Medicine Physician

[Clinic/Hospital Name]

[Clinic Address]

[City, State, ZIP Code]

[Date]

**Neurology Consultant**

[Neurology Clinic Name]

[Neurologist’s Address]

[City, State, ZIP Code]

**Re: Neurology Referral for Transient Ischemic Attack Evaluation**

**Patient Name:** [Patient’s Full Name]

**Date of Birth:** [Patient’s DOB]

Dear Dr. [Neurologist’s Last Name],

I am referring Mr. [Patient’s Last Name], a 68-year-old right-handed male, for evaluation of a suspected **transient ischemic attack (TIA)** that occurred earlier today. Below is a summary of his presentation and findings:

#### History of Present Illness

Mr. [Last Name] experienced sudden-onset **left-sided weakness and difficulty speaking** approximately **30 minutes** before arrival at my clinic. He described numbness and tingling in his left arm and leg, as well as **slurred speech and word-finding difficulty**. These symptoms **fully resolved within 15 minutes**. He denies headache, dizziness, vision changes, or loss of consciousness.

His past medical history includes **hypertension, hyperlipidemia, and type 2 diabetes mellitus**. He underwent **coronary artery bypass grafting in 2018**. He is a **retired construction worker**, a **former smoker (quit 10 years ago),** and an **occasional social alcohol drinker**.

#### Physical Examination

- **General:** Well-nourished, alert, and oriented male in no acute distress.
- **Vital Signs:** BP 150/95 mmHg, HR 78 bpm, RR 16, BMI 27.
- **Cardiac & Pulmonary:** Regular rate and rhythm; clear to auscultation.
- **Neurological Exam: Normal on arrival**; no motor or sensory deficits.

#### Initial Workup & Differential Diagnosis

**Primary Diagnosis:** Suspected TIA. Given his risk factors (hypertension, diabetes, hyperlipidemia, smoking history), his symptoms warrant further evaluation.

##### Differential Diagnoses

- Stroke (though complete symptom resolution suggests TIA)
- Migraine aura without headache
- Hypoglycemia (unlikely given normal blood glucose)

#### Plan & Referral Request

I am requesting **urgent neurology consultation** to further assess stroke risk and management. Workup and next steps include:

- **Immediate imaging: Non-contrast head CT** to rule out hemorrhage
- **Vascular studies: Carotid ultrasound** for stenosis evaluation
- **Cardiac assessment: Echocardiogram, ECG** to assess embolic sources
- **Risk factor management: Initiation of aspirin/clopidogrel, blood pressure control**

Your expertise in determining **long-term secondary prevention strategies** and further workup (e.g., MRI brain, MRA) would be highly valuable. Please let me know if additional information is needed.

Thank you for your evaluation and guidance.

**Sincerely,**

[Your Full Name, MD]

[Your Contact Information]

### A.2: Low-Quality Referral Letter (Radiculopathy Case – Letter 5.2, Score: 10/30)

**[Referring Physician’s Name]**

Family Medicine Physician

[Clinic/Hospital Name]

[Clinic Address]

[City, State, ZIP Code] [Date]

**Neurology Consultant**

[Neurology Clinic Name]

[Neurologist’s Address]

[City, State, ZIP Code]

**Re: Neurology Consultation for [Patient’s Name]**

Dear Dr. [Neurologist’s Last Name],

I am referring **[Patient’s Name], a [Age]-year-old [Gender]**, for further neurological evaluation. He presents with **[brief summary of symptoms]**, which have persisted for **[duration]**.

#### Clinical History

- **Presenting Complaint: [Details of symptoms, onset, progression, severity, any related factors]**
- **Past Medical History: [Relevant conditions, surgeries, or major illnesses]**
- **Medications: [Current medications, dosages, recent changes]**
- **Family History: [Pertinent neurological or other conditions]**

#### Physical Examination

- **General Appearance: [Patient’s general condition, distress, etc.]**
- **Neurological Examination: [Summary of findings: mental status, cranial nerves, motor function, reflexes, gait]**

#### Investigations & Consultation Request

**[List any prior workup performed, e.g., imaging, labs, pending tests]**

I am requesting **[neurologist’s input regarding differential diagnoses, further workup, and management options]**.

Please let me know if you need any further details. Thank you for your assessment and recommendations.

**Sincerely,**

[Your Full Name, MD]

[Your Contact Information]

## Appendix A Commentary

- Letter 3.10 demonstrates strong organization, completeness, and clinical clarity, scoring 30/30.
- Letter 5.2 lacks key details in history, physical examination, and plan, as the LLM left the information areas incomplete (10/30).

These examples highlight the variability in AI-generated referral letters and emphasize the importance of physician oversight in clinical documentation.

